# Positive association of Angiotensin II Receptor Blockers, not Angiotensin-Converting Enzyme Inhibitors, with an increased vulnerability to SARS-CoV-2 infection in patients hospitalized for suspected COVID-19 pneumonia

**DOI:** 10.1101/2020.08.30.20182451

**Authors:** Jean-Louis Georges, Floriane Gilles, Hélène Cochet, Alisson Bertrand, Marie De Tournemire, Victorien Monguillon, Maeva Pasqualini, Alix Prevot, Guillaume Roger, Joseph Saba, Joséphine Soltani, Mehrsa Koukabi-Fradelizi, Jean-Paul Beressi, Cécile Laureana, Jean-François Prost, Bernard Livarek

**Affiliations:** Department of cardiology; Emergency department; Department of diabetology; Department of medical information, Centre Hospitalier de Versailles, Le Chesnay, France

**Keywords:** COVID-19, Angiotensin-Converting Enzyme Inhibitors, Angiotensin II Receptor Blockers, Renin-Angiotensin System, Hypertension

## Abstract

**Background:** Angiotensin converting enzyme (ACE) type 2 is the receptor of SARS-CoV-2 for entry into lungs cells. Because ACE-2 may be modulated by ACE inhibitors (ACEIs) and angiotensin II receptor blockers (ARBs), there is concern that patients treated with ACEIs and ARBs are at higher risk for COVID-19 infection.

**Aim:** This study sought to analyze the association of COVID-19 with previous treatment with ACEI and ARB.

**Methods:** We retrospectively reviewed 684 consecutive patients hospitalized for suspected COVID-19 pneumonia and tested by PCR. Patients were split into 2 groups, whether (group 1, n=484) or not (group 2, n=250) COVID-19 was confirmed. Multivariate adjusted comparisons included a propensity score analysis.

**Results:** Age was 63.6±18.7 years, and 302(44%) were female. Hypertension was present in 42.6% and 38.4% patients of group 1 and 2, respectively (P=0.28). A treatment with ARBs (20.7% versus 12.0%, respectively, OR 1.92, 95% confidence interval [1.23-2.98], p=0.004) was more frequent in patients of group 1 than in group 2. No difference was found for treatment with ACEIs (12.7% vs 15.7%, respectively, OR 0.81 [0.52-1.26], p=0.35). Propensity score matched multivariate logistic regression confirmed a significant association between COVID-19 and a previous treatment with ARBs (adjusted OR 2.18 [1.29-3.67], p=0.004). Significant interaction between ARBs and ACEIs for the risk of COVID-19 was observed in patients aged>60, women, and hypertensive patients.

**Conclusion:** This study suggests that ACEIs and ARBs are not similarly associated with the COVID-19. In this retrospective series, patients with COVID-19 pneumonia received more frequently a previous treatment with ARBs, than patients without COVID-19.

## INTRODUCTION

The coronavirus disease 2019 (COVID-19), caused by the severe acute respiratory syndrome coronavirus 2 (SARS-CoV-2), had been officially declared by the World Health Organization as a global pandemic on March 11, 2020, and had been the main challenge that health care providers have had to face. The relationships between COVID-19 and the renin-angiotensin-aldosterone system and its inhibitors have been widely debated. SARS-CoV-2 uses the angiotensin-converting enzyme 2 (ACE-2) as a cellular entry receptor **(1, 2)**. ACE-2 is furthermore a key enzyme of the renin-angiotensin-aldosterone system (RAAS) that is likely to be modulated by the use of either angiotensin converting enzyme inhibitors (ACEIs) or angiotensin II type 1 (AT1) receptor blockers (ARBs) **(3, 4)**. ACE-2 may have a protective effect against lung injury because it degrades angiotensin II to angiotensin-(1-7) **(5)**. The effect of RAAS inhibition on ACE-2 expression is complex **(4, 6, 7)**, and poorly studied in humans **(8, 9)**.

ARBs have been demonstrated to be protective against lung injury in different experimental models of acute respiratory distress syndrome (ARDS), infective or not **(5, 10-12)**. In hypertensive patients already affected by COVID-19 pneumonia, there is accumulating evidence that ACEI/ARB treatment is associated with a lower mortality **(13, 14)**.

In contrast, there is no evidence that ACEIs and ARBs could affect patients’ vulnerability to COVID-19. A study conducted in a large US population found no association between ACEI or ARB use and COVID-19 test positivity, although hospitalizations related to COVID-19 were more frequent in patients treated by ACEIs/ARBs **(15)**. Another case control study found a positive association between RAAS blockers and the risk of COVID-19 that was explained by a higher prevalence of cardiovascular disease **(16)**. Two other case-control studies were negative **(17, 18)** and showed no difference between ACEIs and ARBs.

However, ACEIs and ARBs have different effect on the RAAS **(4, 6)** as well as on the risk of non COVID-19 pneumonia **(19)**. Their interaction with COVID-19 may therefore differ, with the hypothesis that ACEIs could be more protective than ARBs against infection.

This study sought to compare the prevalence of hypertension and previous treatments with ACEIs and ARBs at admission in a consecutive series of high-risk patients suspected of COVID-19 acute pneumonia, hospitalized for confirmation or not of COVID-19 in a tertiary center located in the Greater Paris area – one of the region most affected by COVID-19 in France.

## METHODS

### Study Design

The COVHYP study is a retrospective observational study that was prospectively planned in March, 2020 at the beginning of the COVID-19 outbreak in the Greater Paris Area in France, and registered in May 2020 (NCT 04374695). The Centre Hospitalier de Versailles is a tertiary hospital that serves a population of about 600.000 inhabitants. The study was conducted in accordance with the principles of the Declaration of Helsinki and protocol was approved by a national committee of research and the French “Commission Nationale Informatique et Libertés”. Patients and/or legal representatives received informed consent. Analyses were retrospective.

### Study Population

From March 10 to April 15, 2020, all consecutive patients referred to the emergency department and hospitalized in a temporary 24-72 hours “COVID-19 screening hospitalization unit” were screened for inclusion. According to regional governmental guidelines, hospitalization was required for patients suspected of COVID-19 having at least one criteria of severity (respiratory frequency >22/min, spontaneous SpO2 <90%, systolic blood pressure <90 mmHg, alteration of consciousness, fast worsening of the general status or serious dehydration in the elderly), or having no severity criteria but a medical history or comorbidities known to increase the risk in case of COVID-19 (listed in supplementary material).

Patients were included into the study if they fulfilled the additional criteria as follows: 1) age ≥18 years; 2) Clinical presentation suggestive of COVID-19 pneumonia, at least: fever > 38°C or influenza-like symptoms (deep asthenia, myalgia, chills, muscular aches) associated with cough or dyspnea or need for oxygen supply (SpO2 ≤90%); and 3) Test of the presence of the SARS-CoV-2 by reverse transcriptase polymerase chain reaction (RT-PCR) in nasopharyngeal or sputum samples. Exclusion criteria were the absence of clinical symptoms of COVID-19, no PCR performed, age < 18 year, prisoners or detainees, and refusal to participate.

Laboratory confirmation for SARS-CoV-2 was defined as a positive result of real-time reverse transcriptase–polymerase chain reaction (RT-PCR) assay of nasal and pharyngeal swabs, according to the French National Reference Center of Respiratory Viruses and the WHO guidance **(20)**. As appropriate, a second RT-PCR assay from sputum or lower respiratory tract aspirates was proposed when the clinical/radiological probability of COVID-19 was high and the first RT-PCR was negative in swab. Almost all patients underwent a chest x ray imaging by chest radiography and/or chest CT scan at the emergency unit. Antihypertensive and cardiac treatments received prior to admission were not discontinued during the hospitalization in COVID-19 screening hospitalization unit.

### Definition of groups

Patients were split into 2 groups, according to the result of SARS-CoV-2 PCR, chest imaging, and the clinical presentation at discharge from the “COVID-19 screening hospitalization unit”. Group 1 (COVID-19) consisted of patients with positive COVID-19 PCR (confirmed) and patients with symptoms and chest CT-scan abnormalities very likely of COVID-19 despite negative PCR (probable). Group 2 (no COVID-19) included patients with negative PCR and chest imaging not suggestive of COVID-19.

### Data Collection

Clinical, radiological, and laboratory data reported in this study were collected from hospital medical reports. The recorded data included the following: age, sex, initial symptoms, time from first symptoms suggestive of COVID-19 and admission, chest imaging performed, result of RT-PCR, history of hypertension, long-term treatments for hypertension, congestive heart failure, ischemic cardiopathy, RAAS inhibitors (mineralocorticoid receptor blockers (MRBs), ACEIs, ARBs), and medical comorbidities (asthma, chronic obstructive pulmonary disease, other chronic pulmonary diseases, chronic cardiac diseases, cancer, hypothyroidism, allergies, and immunosuppression).

### Statistical Analyses

Continuous data are presented as mean ± standard deviation (SD) or median [interquartile ranges, IQRs] as appropriate, and were compared between groups using analysis of variance or nonparametric Mann-Whitney U test. Categorical variables are presented as counts and percentages, and were compared using the χ^2^ test or the Fischer’s exact test. Multivariate analyses were performed using logistic regression, with adjustment on age, sex, obesity (body mass index >30 kg/m^2^), hypertension and history of chronic cardiac disease.

In addition to the main analysis, as in the observational studies, treatment selection is often influenced by subject characteristics; in order to address the issues of confounding by indication, we used a propensity score-matching analysis to balance the different RAAS treatments groups on the possible baseline confounders Multivariate logistic regressions were performed, and the probability of receiving ARBs (or ACEIs) given the observed covariates was estimated. All the variables listed in the supplementary Table 1 were included in the model regardless of statistical significance.

After fitting the model, patients were ranked by their estimated propensity score and grouped within quintiles. Quintiles are commonly used for adjustment, as they are expected to remove 90% of the confounding. Propensity score adjusted analyses were then performed to compare the association between COVID-19 status and previous treatments, either by univariate analyses by quintiles of propensity score in each group, and multivariate logistic regression including the propensity score as a covariate.

Stratified analyses were performed in pre-specified subgroups according gender, age > 60 years, hypertension, and diabetes, using the Cochran-Mantel-Haenszel Chi^2^ statistics. A *P* value <.05 was considered significant. All statistical analyses were carried out with the SPSS® version 19.0 software (SPSS Inc., Chicago, IL) and the R version i386 3.6.2.software.

## RESULTS

### Baseline and initial symptoms

During the study period, 763 consecutive patients were hospitalized in COVID-19 screening unit, 79 were excluded (supplementary material), and 684 were included in the study. COVID-19 was diagnosed in 434 patients (63.4%, 396 confirmed and 38 probable), and excluded in 250 patients (36.6%). Baseline characteristics of patients of both groups are shown in Table 1. The two groups were well balanced for fever or flu-like symptoms (almost all patients in both groups), cough (69.1% in group 1 vs 65.6% in group 2, respectively), ENT, digestive and neurologic symptoms. Dyspnea (75.8% vs 67.6%, respectively), male gender and time from first symptoms to admission were higher in Group 1 than in group 2. A second RT-PCR in sputum sample has been performed in 55 patients (8.0%) and was positive in 17. Chest CT scan has been performed most frequently in patient with subsequently confirmed COVID-19. A Discrepancy between chest imaging indicated as “suggestive of COVID” by radiologist and discharge diagnosis of “no COVID-19” remained in 7 patients, all with congestive heart failure or chronic pulmonary disease.

**Table 1:**
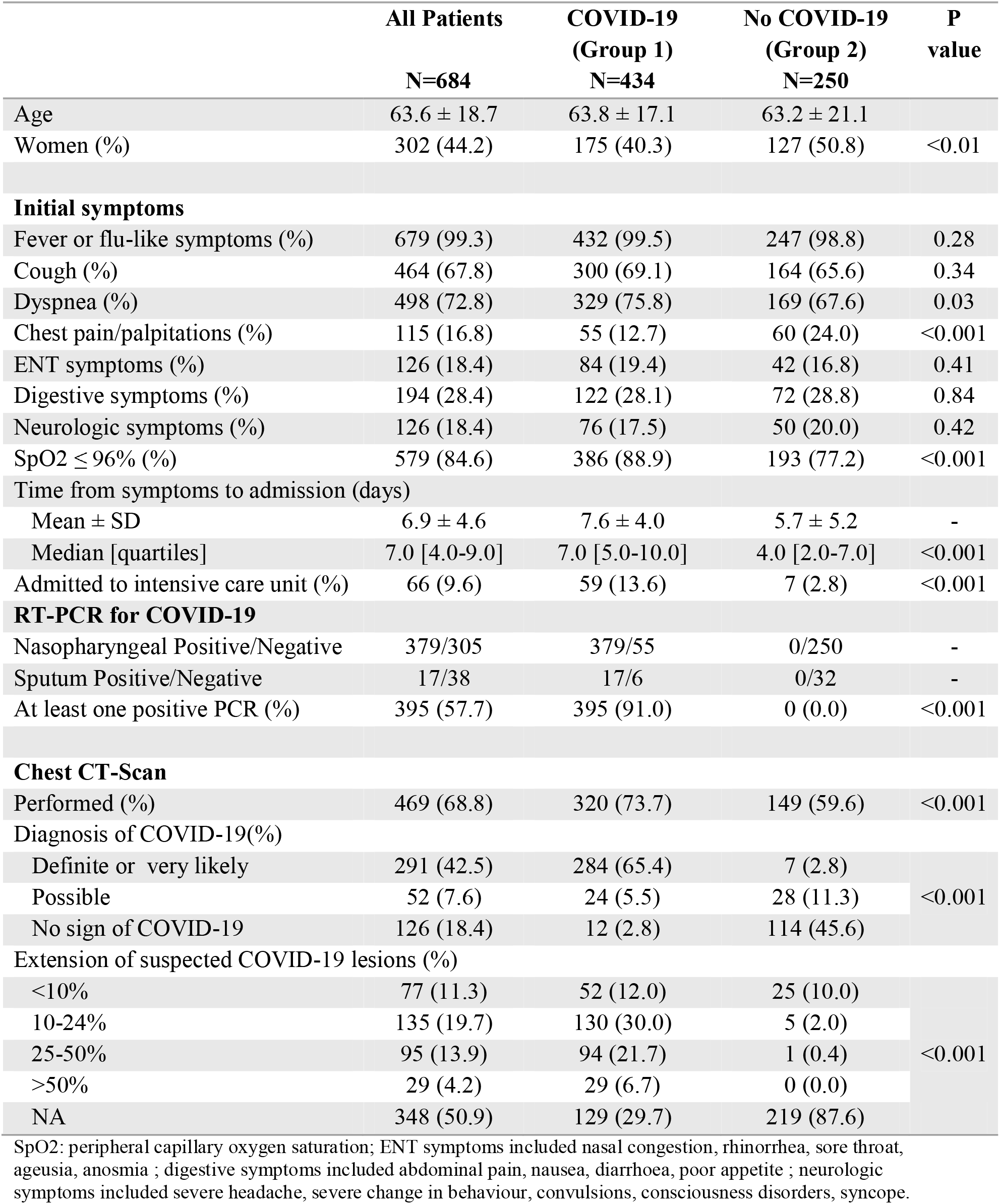
Baseline and admission characteristics.

|  | All Patients | COVID-19<br>(Group 1) | No COVID-19<br>(Group 2) | P<br>value |
| --- | --- | --- | --- | --- |
|  | N=684 | N=434 | N=250 |  |
| Age | 63.6 ± 18.7 | 63.8 ± 17.1 | 63.2 ± 21.1 |  |
| Women (%) | 302 (44.2) | 175 (40.3) | 127 (50.8) | <0.01 |
| Initial symptoms |  |  |  |  |
| Fever or flu-like symptoms (%) | 679 (99.3) | 432 (99.5) | 247 (98.8) | 0.28 |
| Cough (%) | 464 (67.8) | 300 (69.1) | 164 (65.6) | 0.34 |
| Dyspnea (%) | 498 (72.8) | 329 (75.8) | 169 (67.6) | 0.03 |
| Chest pain/palpitations (%) | 115 (16.8) | 55 (12.7) | 60 (24.0) | <0.001 |
| ENT symptoms (%) | 126 (18.4) | 84 (19.4) | 42 (16.8) | 0.41 |
| Digestive symptoms (%) | 194 (28.4) | 122 (28.1) | 72 (28.8) | 0.84 |
| Neurologic symptoms (%) | 126 (18.4) | 76 (17.5) | 50 (20.0) | 0.42 |
| SpO2 ≤ 96% (%) | 579 (84.6) | 386 (88.9) | 193 (77.2) | <0.001 |
| Time from symptoms to admission (days) |  |  |  |  |
| Mean ± SD | 6.9 ± 4.6 | 7.6 ± 4.0 | 5.7 ± 5.2 | - |
| Median [quartiles] | 7.0 [4.0-9.0] | 7.0 [5.0-10.0] | 4.0 [2.0-7.0] | <0.001 |
| Admitted to intensive care unit (%) | 66 (9.6) | 59 (13.6) | 7 (2.8) | <0.001 |
| RT-PCR for COVID-19 |  |  |  |  |
| Nasopharyngeal Positive/Negative | 379/305 | 379/55 | 0/250 | - |
| Sputum Positive/Negative | 17/38 | 17/6 | 0/32 | - |
| At least one positive PCR (%) | 395 (57.7) | 395 (91.0) | 0 (0.0) | <0.001 |
| Chest CT-Scan |  |  |  |  |
| Performed (%) | 469 (68.8) | 320 (73.7) | 149 (59.6) | <0.001 |
| Diagnosis of COVID-19(%) |  |  |  |  |
| Definite or very likely | 291 (42.5) | 284 (65.4) | 7 (2.8) | <0.001 |
| Possible | 52 (7.6) | 24 (5.5) | 28 (11.3) |  |
| No sign of COVID-19 | 126 (18.4) | 12 (2.8) | 114 (45.6) |  |
| Extension of suspected COVID-19 lesions (%) |  |  |  |  |
| <10% | 77 (11.3) | 52 (12.0) | 25 (10.0) | <0.001 |
| 10-24% | 135 (19.7) | 130 (30.0) | 5 (2.0) |  |
| 25-50% | 95 (13.9) | 94 (21.7) | 1 (0.4) |  |
| >50% | 29 (4.2) | 29 (6.7) | 0 (0.0) |  |
| NA | 348 (50.9) | 129 (29.7) | 219 (87.6) |  |

SpO<sub>2</sub>: peripheral capillary oxygen saturation; ENT symptoms included nasal congestion, rhinorrhea, sore throat, ageusia, anosmia ; digestive symptoms included abdominal pain, nausea, diarrhoea, poor appetite ; neurologic symptoms included severe headache, severe change in behaviour, convulsions, consciousness disorders, syncope.

### Comorbidities

The distributions of comorbidities are shown in Table 2. In this series of patients, a negative association was found between COVID-19 and asthma, chronic obstructive pulmonary disease, and chronic heart disease. Non-significant trend for a positive association was found for obesity and hypothyroidism. History of congestive heart failure or left ventricular ejection fraction <40% was present in only 3.2% of patients (2.4% in group 1).

**Table 2:** Comorbidities.

|  | **All Patients** | **COVID-19**<br>**(Group 1)** | **No COVID-19**<br>**(Group 2)** | **P**<br>**value** |
| --- | --- | --- | --- | --- |
|  | **N=684** | **N=434** | **N=250** |  |
| Asthma | 74 (10.8) | 37 (8.5) | 38 (15.2) | <0.01 |
| Chronic pulmonary disease | 61 (8.9) | 30 (6.9) | 31 (12.4) | 0.02 |
| COPD | 50 (7.3) | 24 (5.5) | 26 (10.4) | - |
| CRPD and others | 11 (1.6) | 6 (1.4) | 5 (2.0) | - |
| Sleep Apnea Syndrome | 30 (4.4) | 18 (4.1) | 12 (4.8) | 0.69 |
| Diabetes mellitus | 115 (16.8) | 77 (17.6) | 38 (15.2) | 0.39 |
| Type 1 | 2 (0.3) | 2 (0.5) | 0 (0.0) | - |
| Type 2, oral treatment | 89 (13.0) | 59 (13.6) | 30 (12.0) | - |
| Type 2, insulin | 24 (3.5) | 16 (3.7) | 8 (3.2) | - |
| Obesity | 79 (11.5) | 58 (13.4) | 21 (8.4) | 0.05 |
| Hypertension | 281 (41.1) | 185 (42.6) | 96 (38.4) | 0.28 |
| Chronic heart disease | 170 (24.8) | 82 (18.9) | 64 (25.6) | 0.04 |
| Coronary artery disease | 53 (7.8) | 31 (7.1) | 22 (8.8) | 0.43 |
| Dilated cardiomyopathy | 8 (1.2) | 3 (0.7) | 5 (2.0) | 0.13 |
| Hypertrophic cardiomyopathy | 3 (0.4) | 2 (0.5) | 1 (0.4) | - |
| Valvular heart disease | 21 (3.1) | 15 (3.4) | 6 (2.4) | 0.45 |
| Arrhythmias | 85 (12.4) | 44 (10.1) | 41 (16.4) | 0.02 |
| Congestive heart failure | 22 (3.2) | 10 (2.3) | 12 (4.8) | 0.14 |
| History of cancer | 106 (15.5) | 62 (14.3) | 44 (17.6) | 0.25 |
| Immunosuppression | 50 (7.3) | 28 (6.5) | 22 (8.8) | 0.26 |
| Allergies | 77 (11.3) | 51 (11.8) | 26 (10.4) | 0.59 |
| Hypothyroidism | 57 (8.3) | 43 (9.9) | 15 (6.0) | 0.08 |

COPD: chronic obstructive pulmonary disease; CRPD: chronic restrictive pulmonary disease

### Hypertension and RAAS inhibitors

Hypertension was present in 42.6% and 38.4% patients of group 1 and 2, respectively (P=0.28) (Table 1), and increased with age, without differences between groups (Figure 1). Distribution of RAAS inhibitors in both groups are shown in Table 3. No patient received the association valsartan-sacubitril, and 1 patient received both an ACEI and an ARB. At least one inhibitor of the RAAS (ACEI, ARB or MRB) was given in 34.1% of patients of group 1 and 26.8% of patient of group 2 (OR 1.41, 95% confidence interval [1.00 – 1.99], p=0.05). Patients of group 1 received more frequently a treatment with ARB as compared to those of group 2 (20.7% versus 12.0%, respectively, OR 1.92, 95%CI, [1.23 – 2.98], p=0.004]). No difference was found for ACEIs (12.7% vs 15.7%, respectively, OR 0.81, 95%CI, [0.52 – 1.26], p=0.35) (Figure 2). These trends, although not significant, were also observed in the subgroup of hypertensive patients (Table 3), who received more frequently an inhibitor of the RAAS in group 1 and other classes of antihypertensive agents in group 2.

**Figure 1:**
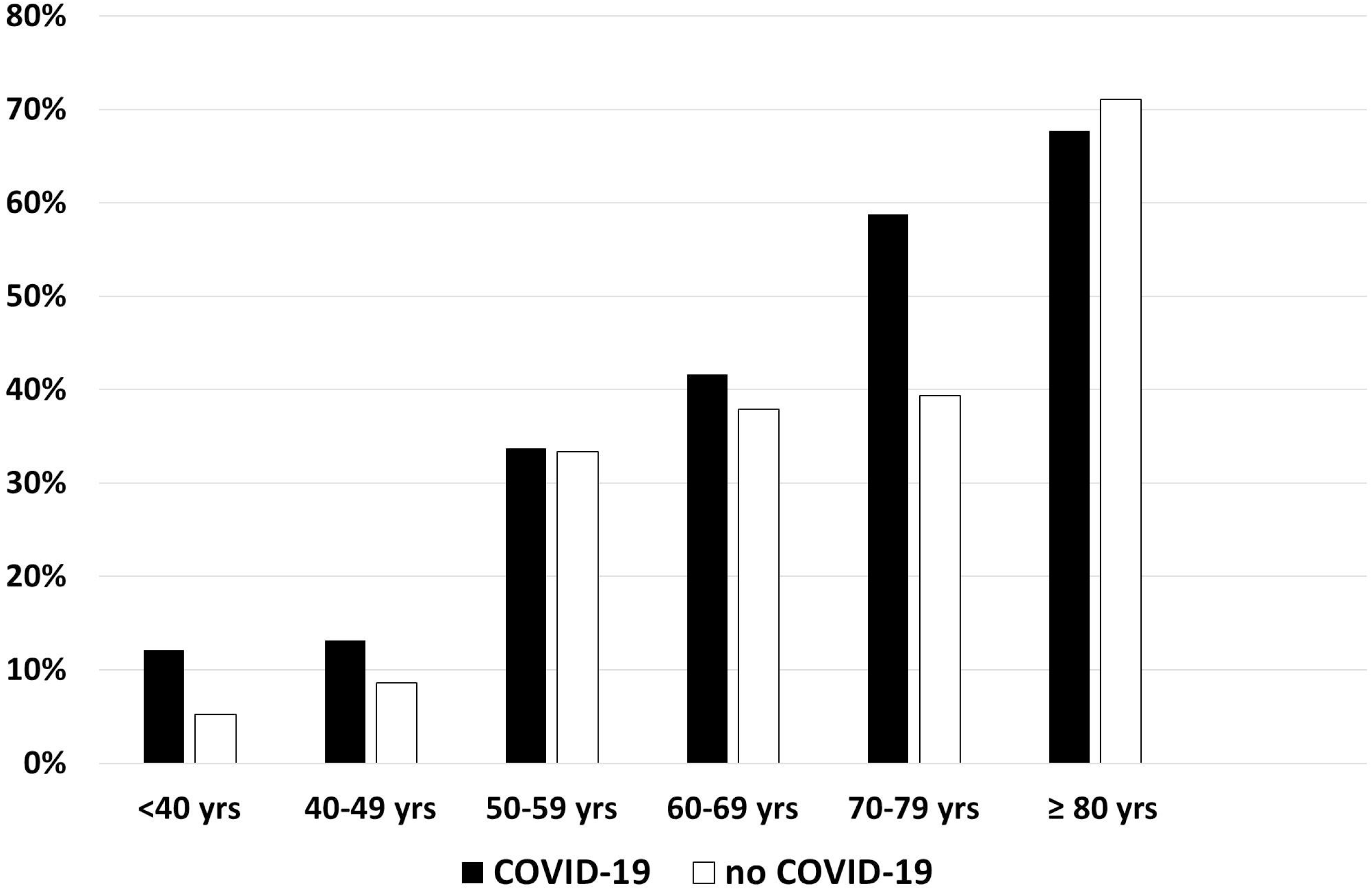
Percentage of hypertension by classes of age and COVID-19 status.

**Figure 2:**
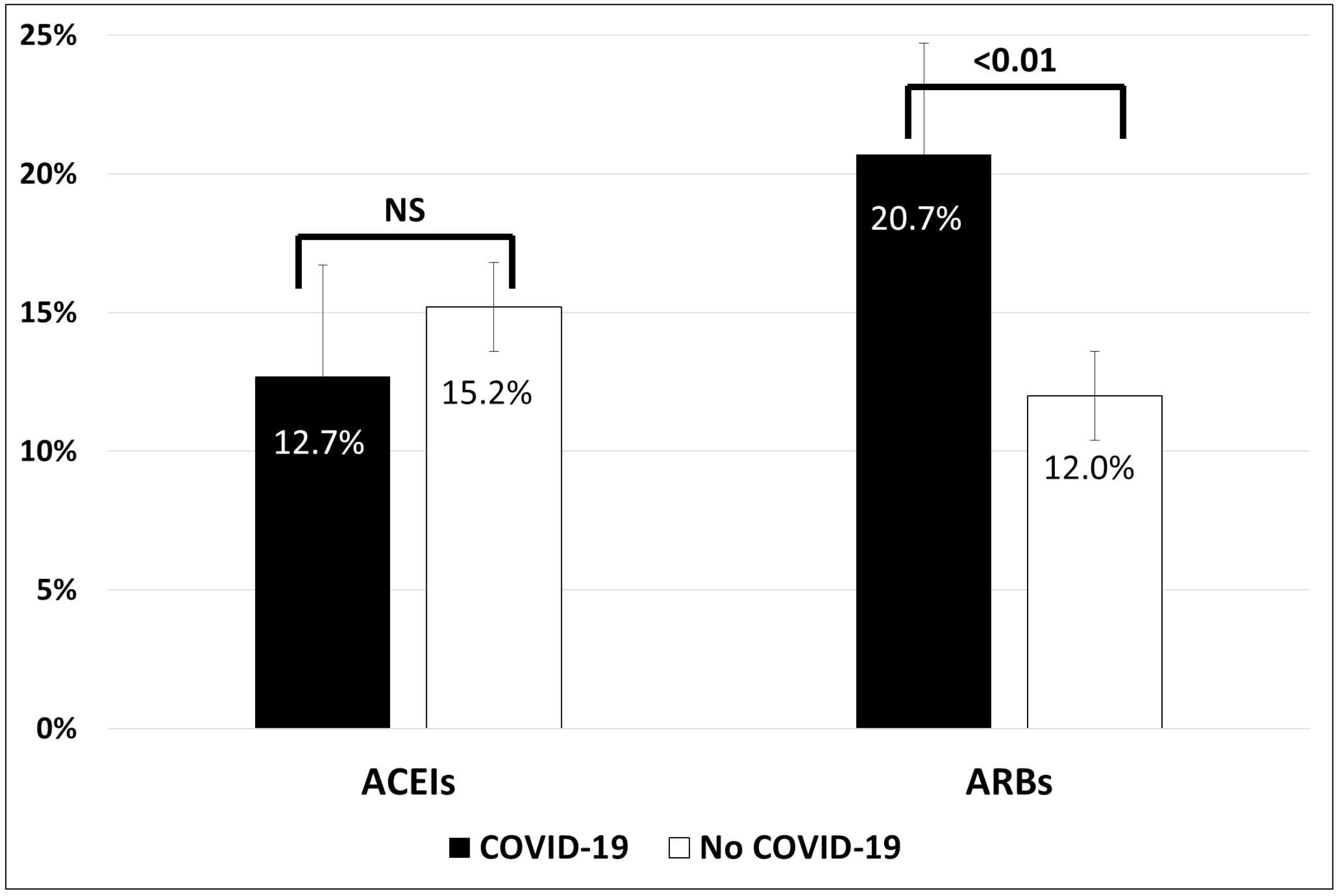
Prevalence of previous treatment with angiotensin converting enzyme inhibitors (ACEIs) and angiotensin II type 1 receptor blockers (ARBs) in patients with and without COVID-19

**Table 3:** Association between previous treatment by RAAS antagonists and COVID-19.

|  | All Patients<br>N=684 | COVID-19<br>(Group 1)<br>N=434 | No COVID-19<br>(Group 2)<br>N=250 | Odds Ratio<br>[95%CI] | P<br>value |
| --- | --- | --- | --- | --- | --- |
| **RAAS inhibitors** |  |  |  |  |  |
| ACEI | 93 (13.6) | 55 (12.7) | 38 (15.2) | 0.81 [0.52-1.23] | 0.35 |
| ARB | 120 (17.5) | 90 (20.7) | 30 (12.0) | 1.92 [1.23-2.98] | 0.004 |
| MRB | 6 (0.9) | 6 (1.4) | 0 (0.0) | - | 0.06 |
| ≥ 1 RAAS inhibitor* | 215 (31.4) | 148 (34.1) | 67 (26.8) | 1.41 [1.00-1.99] | 0.05 |
| **Indication for RAAS inhibitors** |  |  |  |  |  |
| Hypertension | 203 (29.7) | 140 (32.2) | 63 (25.2) | - | 0.02 |
| Congestive heart failure | 5 (0.7) | 3 (0.7) | 2 (0.8) | - | 0.96 |
| Ischaemic cardiopathy | 21 (3.1) | 13 (3.0) | 8 (3.2) | - | 0.90 |
| **Patients with hypertension** |  |  |  |  |  |
|  | N=281 | N=185 | N=96 |  |  |
| ACEI | 83 (29.5) | 48 (25.9) | 35 (36.5) | 0.61 [0.36-1.04] | 0.07 |
| ARB | 118 (42.0) | 89 (48.1) | 29 (30.2) | 2.14 [1.28-3.59] | 0.004 |
| ≥ 1 RAAS inhibitor* | 203 (72.2) | 140 (75.7) | 63 (65.6) | 1.63 [0.95-2.79] | 0.08 |
| Other antihypertensive drugs** | 59 (21.0) | 33 (17.8) | 26 (27.1) | 0.58 [0.33-1.05] | 0.07 |
| No antihypertensive drugs | 19 (6.8) | 12 (6.5) | 7 (7.3) | 0.88 [0.34-2.32] | 0.80 |

RAAS: renin angiotensin aldosterone system; ACEI: angiotensin converting enzyme inhibitor; ARB: angiotensin II type 1 receptor blocker; MRB: mineralocorticoid receptor blocker;
\* Totals are not equal to the sums of components, due to associations of RAAS antagonists or multiple indications for RAAS antagonists;
\*\* Treatments with beta-blockers, calcium channel inhibitors, diuretics others than RAAS antagonists

Propensity score matched multivariate logistic regression confirmed a significant association between COVID-19 and a previous treatment with ARBs (adjusted OR 2.18, 95%CI, [1.29 – 3.67], p=0.004) (Supplementary Table 1). Similar results were found when the 396 patients with PCR confirmed COVID-19 were compared to the 250 patients without COVID-19 (supplementary material). Stratified analyses (Figure 3) showed opposite Odds Ratios for the risk of COVID-19 associated with previous ARBs and ACEIs, with significant interactions in women, patients aged > 60 years or with hypertension, and a non-significant trend in diabetic patients.

**Figure 3:**
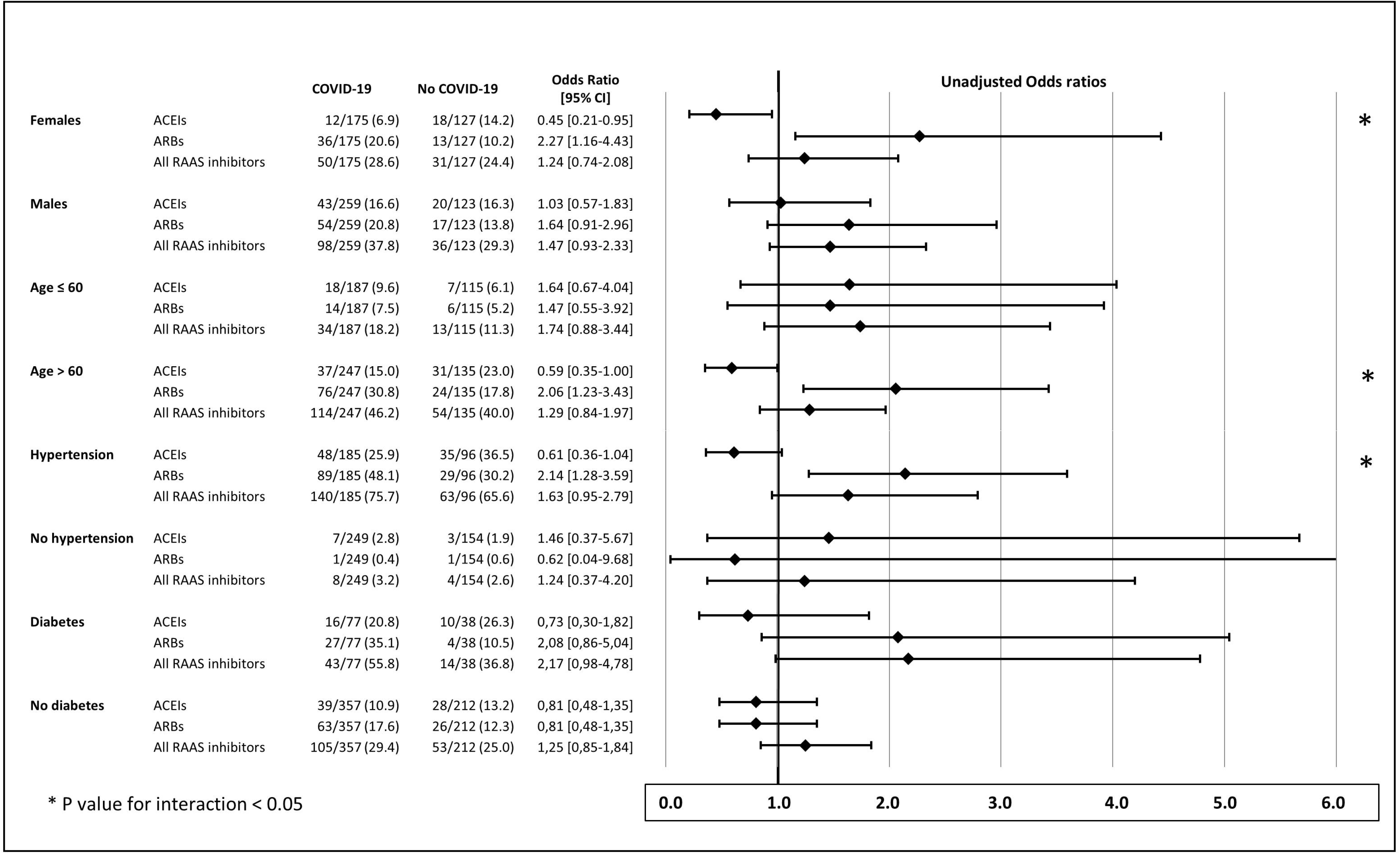
Stratified analysis of relationships between previous treatment with renin angiotensin aldosterone blockers and COVID-19, according to gender, age, hypertension, and diabetes (RAAS: renin angiotensin aldosterone system; ACEIs: angiotensin converting enzyme inhibitors; ARBs: angiotensin II type 1 receptor blockers)

## DISCUSSION

The results of this observational study conducted on a consecutive series of patients with clinical presentation consistent with COVID-19 pneumonia showed a positive association between COVID-19 and a previous treatment with ARBs, and no association with ACEIs. They suggest that a long-term treatment with an inhibitor of RAAS may be not neutral for the vulnerability to the SARS-CoV-2, with a trend for opposite effects of ARBs and ACEIs, particularly in aged patients and women. This may be of importance, since it has been shown that women with hypertension are less frequently treated by ACEIs and ARBs than men **(21)**.

Unfortunately, the present results were discrepant with those of large observational retrospective studies conducted in USA **(15, 17)**, Italy **(16)**, and Denmark **(18)**, which found no difference in the prevalence of RAAS inhibitors between COVID-19 patients and controls. These studies strongly differ from the present study by the selection of either patients or controls. In these studies, a minority of patients tested positive for COVID-19 were hospitalized (25-50%) **(15, 16)**, and the rate of hospitalization in controls is not given. Interestingly, in one study **(15)** a significant association was found between ACEI/ARB treatment and hospitalization, with an odds ratio (1.93, 95%CI, 1.38-2.71) close to that found for ARBs in the present study. For two studies, age- and sex-matched controls were drawn from general population databases, and were not specifically tested for COVID-19 **(16, 18)**. The prevalence of treatment with RAAS inhibitors varied considerably in the overall study populations, from 12.5% and 18.4% In the US studies **(15,17)** to 45.6% in the Italian study **(16)**, and >60% in the Danish study, which was restricted to hypertensive patients **(18)**. In all these studies, baseline characteristics and comorbidities were different in cases and controls. In the present study, both patients and controls were patients who presented to emergency hospital department for symptoms suggestive of acute pulmonary infection, and who were admitted because of severity criteria, including the need for oxygen supply. The more selective inclusion criteria resulted in baseline characteristics, clinical symptoms, and comorbidities relatively well balanced between groups despite the absence of randomization, with the exception that COVID-19 patients had a more severe respiratory presentation (more dyspnea, lower SpO2, higher extension of pulmonary lesions on CT scan, more admission in ICU) than controls. These differences in selection criteria may in part explain the difference in the results.

There are theoretical arguments for different effects of ACEIs and ARBs on the RAAS, and the vulnerability against pulmonary infection. Both ACEI and ARB have been shown to increase cardiac ACE-2 gene transcription in some animal models **(3, 5)**, but there is no evidence that inhibitors of the RAAS upregulate the transmembrane ACE-2 receptor expression in human lung **(22)**. Moreover, several experimental and clinical data suggest that ACEIs and ARBs have not similar effects on ACE-2 expression and activity. In a murine model of myocardial ischemia, the upregulation of ACE-2 induced by lisinopril was higher than that induced by losartan, but was associated with no increase in cardiac ACE-2 activity. In the same model, lisinopril and losartan were associated with opposite variations in plasma Angiotensin (Ang) II and Ang(1-7)/Ang II ratio **(4)**. Conflicting evidence was also reported with ramipril that failed to increase ACE-2 **(8)**. Discrepant effects of ACEIs and ARBs on ACE-2 mRNA and activity, as well as RAAS metabolism have been summarized by Kreutz et al. **(6)**.

Differences between ACEI and ARBs have also been found for the human vulnerability to pulmonary infections. In a meta-analysis of 37 studies **(17)**, ACEIs were associated with a reduced risk of pneumonia as compared with control treatment (OR 0.66, 95%CI, 0.55 to 0.80), but also with ARBs (OR estimate 0.69, 95%CI, 0.56 to 0.85). In contrast, similar protective effects of either ACEIs or ARBs on the risk of influenza have been demonstrated in a large British population **(23)**. During a median 8.7 years of follow-up, patients who were treated with ACEIs had a lower risk of influenza than those who had not (adjusted hazard ratio 0.66; 95%CI, 0.62 to 0.70). Conversely, in-hospital usage of ACEIs trended to have a higher incidence and risk of 28-day COVID-19 mortality than usage of ARBs in a retrospective Chinese study **(24)**. One of our hypotheses when planning this study was that ACEIs could have a protective effect against the risk of COVID-19, and not ARBs. Although odds ratios for COVID-19 pneumonia associated with ACEIs and ARBs varied inversely in the present study, we did not confirm a protective effect of long-term treatment with ACEIs in all patients. The results suggest a significant association between the risk of developing COVID-19 pneumonia and a previous treatment with ARBs. The protective effect associated with ACEIs was however likely in women and patients aged > 60 years (Figure 3).

This study did not investigate the association between RAAS inhibitors and the severity of COVID-19. This point is of importance since there is accumulating evidence that ARBs have protective effects on the development of ARDS triggered by pathogens, including the Coronaviruses **(10-12)**. Genetic deletion of ACE-2 worsened experimental ARDS **(1)**, while Ang (1-7) and blockade of AT1 receptors improved it **(4)**. In humans, ACEIs and ARBs were neutral **(25)** or were associated with a decrease in severity of pneumonia due to COVID-19 **(13**) and mortality among hypertensive patients **(14**).

It is therefore necessary to clearly distinguish two aspects of the relationship between ACEIs and ARBs, and COVID-19. The first one is the questionable protective or deleterious effect of ACEIs and ARBs on SARS-CoV-2 lung invasion and infection. The second one is the likely protective effect of RAAS inhibitors against progression of COVID-19 and ARDS, when lungs are already infected. The results of the present study are restricted to the vulnerability to SARS-CoV-2 infection, and do not involve the severity of the COVID-19 pneumonia. They suggest a negative effect of ARBs, not of ACEIs. The results have to be confirmed by data from large observational studies or even randomised clinical trials. Until results of confirmatory studies are not available, and because discontinuation of ACEIs or ARBs may be harmful in high-risk patients **(26)**, recommendations for continuing RAAS inhibitors in patients affected by-or at high risk for COVID-19 should be respected **(27)**.

### Study Limitations

This study has several limitations. Although it was prospectively designed, collection and analyses of data were retrospective. The biases classically associated with retrospective studies may account for the observed differences. Particularly, a misclassification of patients with and without COVID-19 may have occurred. In case of discrepancy between typical clinical symptoms/chest CT scan abnormalities and a first negative RT-PCR in swab, routine practice was to perform a second PCR in sputum sample, as appropriate, and reach a final collegial diagnosis with clinicians and radiologists if the second PCR was negative. In our study 38/384 of patients of group 1 were diagnosed having probable COVID-19 despite negative PCR. This corresponds to an overall RT-PCR false negative rate of 10%, much lower than the 30% false negative rate reported in Wuhan, China **(28)**. Conversely, few patients who tested negative for PCR were classified as “no COVID-19”, although abnormalities in chest CT scan were consistent with COVID-19. Most of these latter patients had non typical symptoms and an alternative diagnosis (pulmonary infection complicated congestive heart failure or chronic pulmonary disease). Nevertheless, in order to take into account and overcome this putative bias, additional analyses were done excluding the 38 patients with probable COVID-19 (Supplementary Table 3), and then pooling the probable COVID-19 with the non-COVID-19 patients (Supplementary Table 4). Similar results were found in the first case, and borderline non-significant results in the second case, the least favorable to the hypothesis of a significant association between ARBs and COVID-19.

Because of potential confounding factors, analyses were adjusted on propensity scores taking into account variates that were independently associated with previous treatments by ARBs or ACEIs. Propensity score adjusted analyses confirmed the positive association between ARBs and COVID-19. Finally, individual creatinine levels were not collected and no adjustment was made on renal function, a potential confounding factor.

### Conclusions

This study confirmed that, overall, RAAS blockers are not associated with the risk of COVID-19. However, comparative analyses suggested that ACEIs and ARBs are not similarly associated with the COVID-19 incidence, the patients with COVID-19 pneumonia receiving more frequently a previous treatment with ARBs than patients without COVID-19. An opposite effect of ACEIs, likely to be protective, and ARBs, not protective, was observed in women and aged patients. The results of the present study need to be interpreted with caution, given the retrospective monocentric observational design of the study. These results have to be confirmed and do not question the current recommendations to continue long term treatments with ACEIs and ARBs, particularly in patients already infected by SARS-CoV-2.

## Data Availability

All relevant data are within the manuscript and its Supporting Information files

## Acknowledgments

We thank Stéphanie Marque Juillet, MD, and all the biologists and technicians of the department of virology, Maxime de Malherbe, MD, François Mignon, MD, Pénélope Labauge, MD, and all the radiologists and radiology technicians of the radiology department, the physicians and nurses of emergency department, departments of diabetology, and cardiology of the Centre Hospitalier de Versailles. We thank Jean-Baptiste Azowa and Karelle Aumasson for their technical assistance, and help in the data collection.

## Authors contributions

JLG, MKF, JPB, CL, and BL contributed to the conception or design of the work. JLG, FG, HC, AB, MDT, VM, AP, GR, JSa, JSo, JFP contributed to the acquisition, analysis, or interpretation of data for the work. JLG and GR drafted the manuscript. All authors critically revised the manuscript. All gave final approval and agree to be accountable for all aspects of work ensuring integrity and accuracy.

